# Performance of BD Onclarity HPV assay on FLOQSwabs vaginal self-samples

**DOI:** 10.1101/2023.07.08.23292408

**Authors:** Marianna Martinelli, Ardashel Latsuzbaia, Jesper Bonde, Helle Pedersen, Anna D. Iacobone, Fabio Bottari, Andrea F. Piana, Roberto Pietri, Clementina E. Cocuzza, Marc Arbyn, Extended Valhudes Study Group

**Affiliations:** Department of Medicine and Surgery, University of Milano-Bicocca, Monza, Italy; Unit of Cancer Epidemiology, Belgian Cancer Centre, Sciensano, Brussels, Belgium; Molecular Pathology Laboratory, Department of Pathology, af. 134, Copenhagen University Hospital, AHH-Hvidovre Hospital, Hvidovre, Denmark; Preventive Gynecology Unit, European Institute of Oncology IRCCS, Milan, Italy; Division of Laboratory Medicine, European Institute of Oncology IRCCS, Milan, Italy; Department of Medicine, Surgery and Pharmacy, University of Sassari, Sassari, Italy; U.O. Coordinamento Consultori Familiari, ASSL Sassari – ATS Sardegna, Sassari, Italy; Department of Human Structure and Repair, Faculty of Medicine and Health Sciences, University Ghent, Ghent, Belgium

**Keywords:** Vaginal self-sampling, HPV, cervical cancer prevention

## Abstract

This study assessed the accuracy of hrHPV testing of BD Onclarity™ HPV (Onclarity) assay on vaginal self-collected FLOQSwab® versus cervical samples to ensure similar accuracy to detect cervical intraepithelial neoplasia. Testing was performed on two automated platforms, BD Viper™ and BD COR™, to evaluate the effect of machine and using two vaginal self-samples to analyse the influence of collection, transport, and freezing-unfreezing on the results.

A cervical sample and two self-samples were collected from 300 women. The first collected vaginal and the cervical sample were tested on BD Viper™ and the second swab was frozen and subsequently tested on both automated systems. Test results on vaginal and cervical specimens were considered the index and comparator, respectively; colposcopy and histology were reference standards.

Relative sensitivity for ≥CIN2 on vaginal samples analysed using the three different workflows vs the cervical sample was 1.01 (0.97-1.06), 1.01 (0.97-1.06), and 1.00 (0.95-1.05), respectively.

Relative specificity resulted 0.83 (0.73-0.94), 0.76 (0.67-0.87) and 0.82 (0.73-0.92) for the 1st, 2nd self-collected sample tested on BD VIPER™ and 2nd self-collected sample tested on BD COR™.

Cut-off optimization for HPV positivity defined at Ct ≤38.3 for HPV16, ≤34.2 for HPV18 and ≤31.5 for all other types showed an increased relative specificity with similar sensitivity. No significant difference was observed between self-samples tested with the two platforms and between first and second-collected swabs.

Onclarity assay on FLOQSwab^®^ using both platforms showed similar sensitivity but lower specificity to detect ≥CIN2 compared to cervical samples. By cut-off optimization, non-inferior specificity could be reached.

## Introduction

Self-collected samples for high-risk human papillomavirus (HPV) testing are increasingly being implemented in cervical cancer screening as a strategy to either supplement or substitute clinician collected samples. Self-sampling offers clinic independent access to cervical screening whether it is intended as out-reach to under-screened women, providing cervical cancer screening in remote regions with limited access to health care, or in organised screening empowering women with the choice on preferred method of screening participation. Overall, the aim remains to increase participation in cervical screening (1-3). The optimal cost-effective strategy for distribution of self-collection kits depends on the local setting, region and country. Nevertheless, whether self-collection kits are distributed as “direct mail” to all eligible women (4-6), as an opt-in version where women actively have to request the offer of screening by self-sample (6-8), or in a clinic assisted manner (9, 10), HPV self-sampling has shown to be well accepted.

A recent meta-analysis showed that self-collected samples have a sensitivity and specificity *at par* with clinician collected samples if validated PCR-based assays are used (11). In terms of clinical management, HPV self-sampling has proven to be a strong motivator for otherwise long-term unscreened women to attend a clinician collected follow up sample after an HPV positive self-sample (7), offsetting concerns over potential loss to follow-up after self-sampling. Whereas clinician collected liquid based cytology (LBC) samples allow for assessment of both HPV testing and cells of the cervix, self-collected samples allow for highly precise HPV testing when using quality controlled and quality assured analysis protocols.

Nevertheless, for HPV screening self-collected samples to become a mature technology, it requires laboratory test protocols are continuously developed to the highest validation standards (12). In this respect, an HPV self-sample consist of the sampling swab (device) combined with the resuspension medium on which the analysis for HPV is conducted.

Several studies have already reported a similar accuracy of PCR-based HPV tests conducted on self-samples compared to clinician-collected samples (13, 14). However, formal international consensus validation criteria’s for HPV self-samples has yet to be presented. The recent VALHUDES (VALidation of HUman papillomavirus assays and collection DEvices for HPV testing on Self-samples) protocol allows for the evaluation of the clinical performance of HPV assays in combination with different self-sampling devices and constitute a first approach towards validation consensus (15). The present study is a diagnostic test accuracy study complying to the VALHUDES framework and the study addresses the validation of PCR-based HPV assays in conjuncture with a defined self-sampling device and automated HPV test platforms. We present the validation of the clinical accuracy of HPV self-samples analyzed with the Onclarity assay on both instrument platforms available for this HPV test. The Onclarity assay is validated for use in cervical screening (16-19) and recently also for vaginal self-samples using Evalyn Brush (13), and Colli-Pee urine samples (20).

We evaluated vaginal self-samples collected with FLOQSwab^®^ 5E089N (FLOQSwab^®^) in combination with BD HPV Self Collection Diluent and compared to clinician-taken liquid-based cytology (LBC) samples, to detect high-grade cervical intraepithelial neoplasia (CIN of grade 2 or worse [≥CIN2]). Moreover, this evaluation of Onclarity assay on vaginal self-collected samples included testing on both available instrument platforms for use with the BD Onclarity HPV assay, the BD Viper™ and BD COR™ (17, 21, 22). The BD Viper™ platform is a medium throughput test platform for HPV testing, the BD COR™ is a high throughput platform. The non-inferior performance of the Onclarity assay on cervical screening samples on the two automated test platforms has previously been described (21, 23), but not on self-collected specimens.

## Materials and Methods

### STUDY DESIGN AND SAMPLE COLLECTION

A total of 300 women were enrolled in the study between March 2021 and July 2021 (Figure 1) consulting two Italian Colposcopy Centres, Preventive Gynecology Unit, European Institute of Oncology (Istituto Europeo di Oncologia, IEO) in Milan and U.O. Coordinamento Consultori Familiari, ASSL Sassari-ATS Sardegna in Sassari). Median age of the study participants was 40 (range 25-64, IQR: 32-48). Exclusion criteria include: 1) women younger than 25 years, 2) older than 64 years, 3) hysterectomized women, 4) women with known pregnancy. Ten patients were excluded due to inadequate reference test (biopsy result was unsatisfactory), one cervical sample and three vaginal self-samples (two first-collected and one second-collected) were excluded due to invalid internal control (beta-globin). All women were referred as per previous screening to detected cell abnormality. Informed consent was collected upon consultation. All enrolled women received written instructions on how to perform self-sampling. Each woman supplied two self-collected vaginal swabs (labelled “1st” and “2nd” based on the order of collection) using FLOQSwab® 5E089N (Copan Italia Spa, Brescia, Italy). The samples were provided prior to undergoing colposcopy. As per standard colposcopy procedure, a cervical brush specimen using a Cervex-Brush® (Rovers® Medical Devices, The Netherlands) was collected by a gynaecologist and transferred into 20 ml of PreservCyt LBC medium (Hologic® Inc., Bedford, MA, USA). Colposcopy was performed, and a colposcopy-targeted biopsy was collected as per routine management of women with prior cervical lesions. A total of 181 histologies were collected.

**Figure 1.**
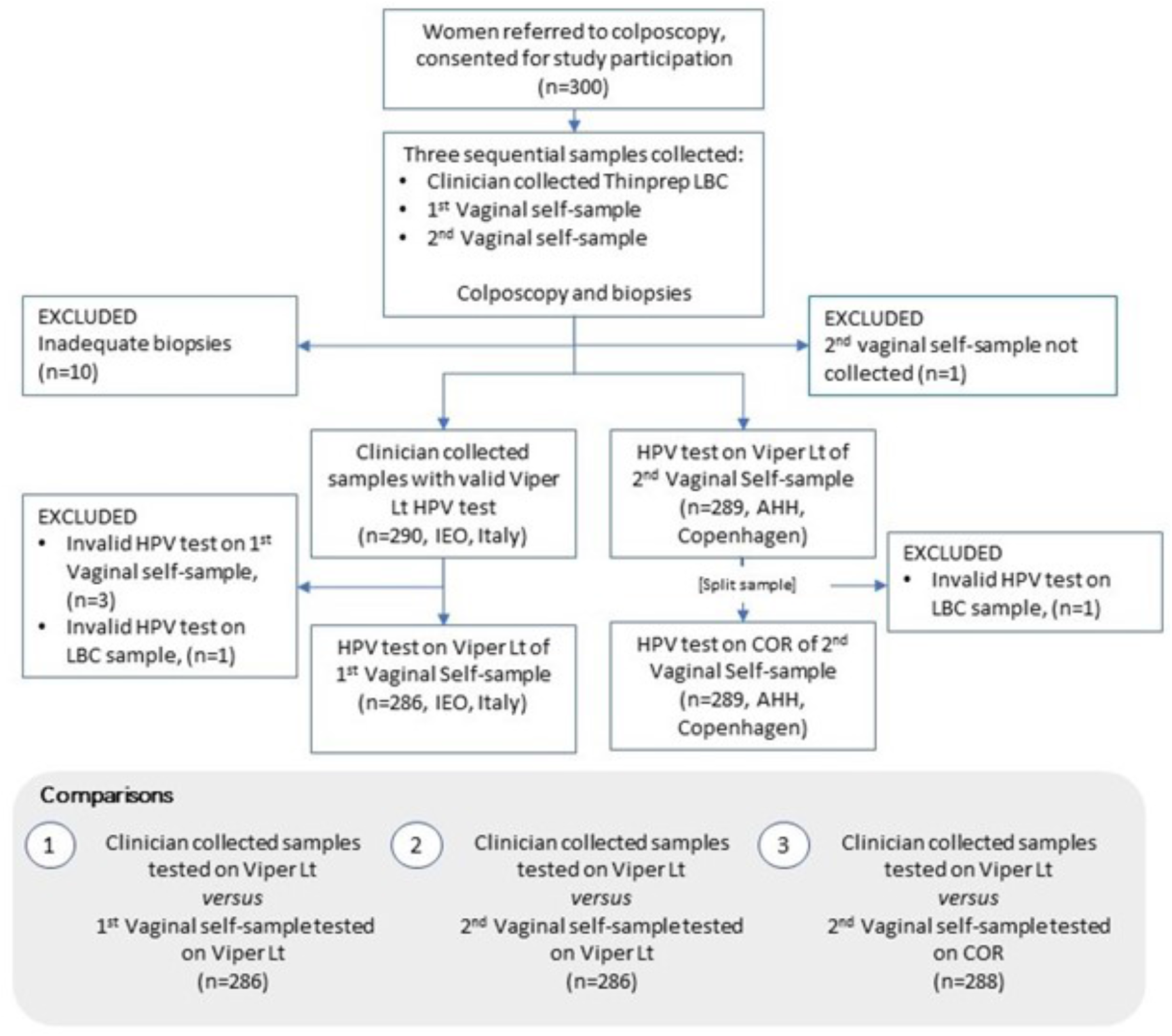
Flow chart of enrolled and tested patients with excluded samples indicated in grey.

### PREANALYTICAL PROCESSING OF SAMPLES

All cervical and vaginal samples collected were labelled with a unique identifier code and sent to the central study laboratory of IEO (Milan, Italy). Cervical samples and dry vaginal self-samples have been stored at room temperature until shipping to the laboratory. Upon arrival in the laboratory, the two FLOQSwabs® were broken into 3 ml tube of medium contained the BD HPV Self Collection Diluent (BD Diagnostics, Sparks, MD, USA). Median interval between first self-sample collection and suspension into BD HPV Self Collection Diluent was 1 day, maximum was 10 days.

The “1st” vaginal self-collected sample was stored at 4°C after resuspension until testing with Onclarity assay on BD Viper™. Median interval between resuspension and testing was 7 days, maximum 18 days.

The “2nd” collected swab was stored at -20°C immediately after broken and resuspension until the end of the study enrolment and subsequently shipped at controlled temperature for analysis at the Molecular Pathology Laboratory, Copenhagen University Hospital, AHH-Hvidovre, Denmark (AHH).

For the second self-sample tested on BD Viper™, frozen samples were thawed immediately before testing.

LBC samples in PreservCyt vial were stored at room temperature (15-30°C) prior to being transferred to a BD HPV LBC diluent tube. Prior to testing, the LBC vial was vortexed for 8-12 seconds followed by immediate transfer of a 0.5 mL aliquot to a BD HPV LBC diluent tube. Tubes were inverted 3-4 times to ensure that the specimen and diluent were well mixed. The remaining volume of the physician-collected sample in the PreservCyt vials was aliquoted at IEO and transferred and stored at the MIRRI (Microbial Resource Research Infrastructure) biobank at the University of Milano-Bicocca (UniMib).

### HPV TESTING

IEO laboratory conducted testing to assess relative clinical sensitivity and specificity on LBC versus the first self-collected sample using Onclarity assay on BD Viper™ platform. AHH used the 2^nd^ self-collected samples to assess the inter-platform accuracy and concordance between the BD COR™ and BD Viper™ systems (Figure 1).

Onclarity assay detects 14 high-risk genotypes and provides the capability of extended genotyping through individual detection of HPV 16, 18, 31, 45, 51, 52 and pooled detection of 33/58, 35/39/68, and 56/59/66 (13, 17, 18, 20, 22, 24). Sample validity control for sample adequacy, sample extraction and amplification efficiency were evaluated by detecting an endogenous human beta-globin sequence. Samples were considered HPV positive if cycle threshold (Ct) value was ≤38.3 for HPV16 and ≤34.2 for all other types, as defined by manufacturer (17). When test failure was reported on one or more sample types, retesting was performed.

### Statistical Analysis

The relative accuracy of BD Onclarity testing on self-samples (index) versus on clinician taken samples (comparator) and 95% confidence intervals were computed taking the matched design into account (15). In addition, we performed direct matched comparisons of first (comparator) with second vaginal (index) self-sample, and second self-sample tested on COR (index) with second self-sample tested on Viper (comparator). Histological outcome and colposcopy results were used as the reference standard. If no biopsy was taken, clinical colposcopy outcome was classified as <CIN2 when colposcopy was satisfactory and did not reveal abnormal findings. In all other cases where a biopsy was performed, the biopsy outcome was used. Post-hoc cut-off optimization (Ct value ≤38.3 for HPV16, ≤34.2 for HPV18 and Ct <=31.5 for others hrHPV) was performed to improve specificity.

The differences in sensitivity and specificity between the specimens was evaluated using McNemar test. Concordance between the specimens was assessed using Cohen’s Kappa (25) and categorized as follows: 0.00 to 0.19 as poor, 0.20 to 0.39 as fair, 0.40 to 0.59 as moderate, 0.60 to 0.79 as good and 0.80 to 1.00 as excellent concordance (26).

We used Wilcoxon signed-rank and Mann-Whitney tests to evaluate the differences in Ct values between specimens. In case of multiple HPV infections, we considered the type with lowest Ct value. Statistical analyses were performed using Stata 16 (College Station, TX, USA).

### Ethical Approval

Extended VALHUDES study (NCT04788849) was approved by the central Ethics Committee of the IEO (IEO 1368, CTO/017-21/MM/gp) and ATS Sardegna (Prot. 258/2020/CE) Ethics Committees. Written informed consent was obtained from all study participants prior to enrolment.

### Data Availability

Final study datasets generated by the study will be stored locally and securely at Sciensano. Anonymized data will be available by request to the corresponding author on a case-by-case basis pending approval from the information security coordinator at Sciensano.

## Results

Characteristics of the study population are present in Table 1. Sixty-four percent of women underwent biopsy or endocervical curettage (181/290) and had subsequent histology evaluation. A total of 207 (207/290, 71.4%) women had ≤CIN1 or a colposcopy without a histological outcome. For women with normal colposcopy and no histology, no disease was assumed. Resulting histology showed 83 women with ≥CIN2 (83/290; 28.6%) including 48 women with ≥CIN3 or worse (48/290; 16.6%).

**Table 1.**
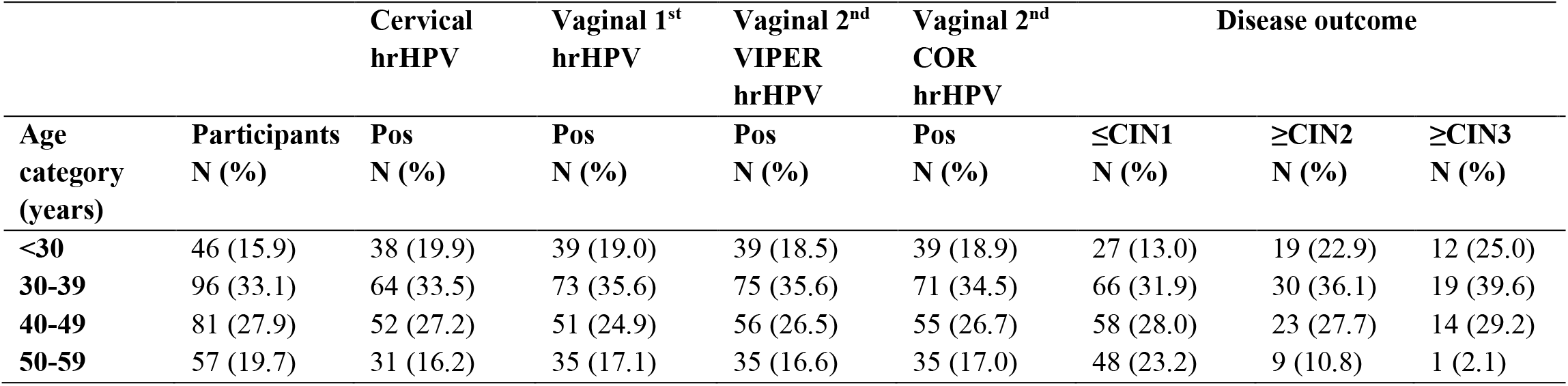
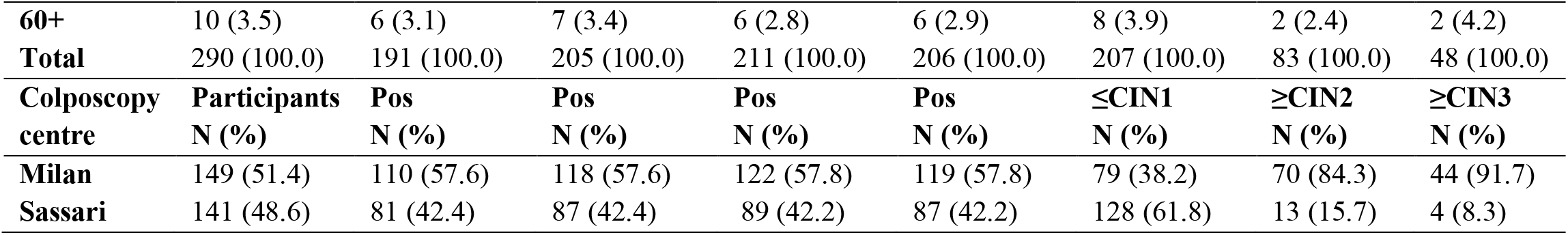
HPV prevalence and disease outcome by age group and colposcopy centres. CIN, cervical intraepithelial neoplasia;

Different denominators were used to evaluate sensitivity and specificity depending on the available valid match samples (Figure 1). Table 2 provides a summary of the data on relative clinical sensitivity and specificity. Data regarding absolute sensitivity and specificity are reported in supplementary material (Table 1S).

**Table 2:** Relative sensitivity for ≥CIN2 and ≥CIN3 and specificity for ≤CIN1 of Onclarity Assay

| Sample type | Relative sensitivity [95%CI] ≥CIN2 | Relative sensitivity [95%CI] ≥CIN3 | Relative specificity [95%CI] ≤CIN1 |
| --- | --- | --- | --- |
| **1<sup>st</sup> vaginal** | 1.01 (0.97-1.06) | 1.00 (0.94-1.06) | 0.83 (0.73-0.94) |
| **2<sup>nd</sup> vaginal (BD Viper™)** | 1.01 (0.97-1.06) | 1.00 (0.94-1.06) | 0.76 (0.67-0.87) |
| **2<sup>nd</sup> vaginal (BD COR™)** | 1.00 (0.95-1.05) | 0.98 (0.90-1.06) | 0.82 (0.73-0.92) |

### Clinical accuracy of BD Onclarity™ HPV Assay

Onclarity assay detected 74 out 83 of ≥CIN2 on cervical samples whereas on the 1st vaginal self-collected samples 75 out 83 of ≥CIN2 cases were detected. Absolute sensitivity was 89.2% (95% CI, 80.4%–94.9%). For the intra-platform reproducibility, paired 2^nd^ self-collected sample showed a clinical sensitivity of 90.2% (74/82; 95% CI, 81.7%–95.7%). HPV testing on the 2nd self-collected sample using BD COR™ detected 74 out 83 cases of ≥CIN2 with a corresponding absolute sensitivity of 89.1% (95% CI, 80.4%–94.9%). The relative clinical sensitivity for ≥CIN2 of Onclarity assay on self-collected samples using BD Viper™ or BD COR™ compared to paired LBC sample was 1.01 (95% CI, 0.97-1.06), 1.01 (95% CI, 0.97-1.06), and 1.00 (95% CI, 0.95-1.05), respectively (Table 2).

Among women with ≤CIN1, 89 were HPV negative on the LBC sample (89/206; 43.2%, 95% CI, 36.3%–50.3%), 74 on the 1^st^ self-collected sample (74/204; 36.3%, 95% CI, 29.7%-43.3%), 68 on 2^nd^ self-collected sample tested with BD Viper™ (68/205; 33.2%, 95% CI, 26.8%-40.1%), and 68 on 2^nd^ vaginal samples tested with BD COR™ (74/206; 35.9%, 95% CI, 29.4%-42.9%) (Table 1S). Compared to LBC, the relative specificity was 0.83 (95% CI, 0.73-0.94), 0.76 (95% CI, 0.67-0.87) and 0.82 (0.73-0.92) for the 1^st^ and 2^nd^ self-collected sample tested on BD Viper™ and 2^nd^ self-collected sample tested on BD COR™, respectively (Table 2).

After cut-off optimization, defined at ≤38.3 for HPV16, ≤34.2 for HPV18 and ≤31.5 cycle thresholds for all other types, an increase of relative specificity was observed with no loss in sensitivity (Supplementary Table S2). No significant difference was observed in the relative sensitivity and specificity between vaginal samples tested with the two platforms (Supplementary Table S3).

### Analytical performance of BD Onclarity™ HPV Assay

Overall and individual genotype concordance between 1st vaginal and cervical sample was moderate, good and excellent with Kappa values between 0.54 and 0.93. Good to excellent concordance was observed between 2nd vaginal sample tested with BD Viper™ and cervical specimen with Kappa values ranging between 0.61 and 0.88. Self-samples tested on BD COR™ showed a moderate to excellent overall and genotype-specific concordance, with Kappa values ranging between 0.52 and 0.91. Data concerning HPV test concordance between cervical and other specimens, overall and by disease status are reported in the supplementary material (Supplementary Tables S4a, b, c, d, e, f). A good agreement in hrHPV detection was observed comparing the results obtained from the 1st and the 2nd self-collected vaginal samples tested using BD Viper™ with a concordance rate of 95.4% (Kappa=0.89). A high percentage of agreement (96.2%, Kappa=0.91) was also demonstrated between 1st self-collected vaginal samples tested using BD VIPER™ and the 2nd self-collected vaginal samples tested using BD COR™. An overall concordance of 97.2% (Kappa=0.93) was found between the results obtain from the analysis of the 2nd vaginal sample tested on both systems.

Overall, median viral Ct values were always significantly higher for cervical compared to vaginal samples (Figure 2 and Supplementary Tables S5a, b, c). No difference was observed in median viral Ct values related to hrHPV detection between first and second vaginal sample tested on BD Viper™ (Supplementary Table 5d). On the contrary, median viral Ct values were lower in 1st vaginal samples tested using BD Viper™ compared to 2nd vaginal samples tested on BD COR™ (Supplementary Table 5e). Similarly, median viral Ct values were lower in 2nd vaginal samples tested using BD Viper™ compared to 2nd vaginal samples tested on BD COR™ (Supplementary Table 5f).

**Figure 2.**
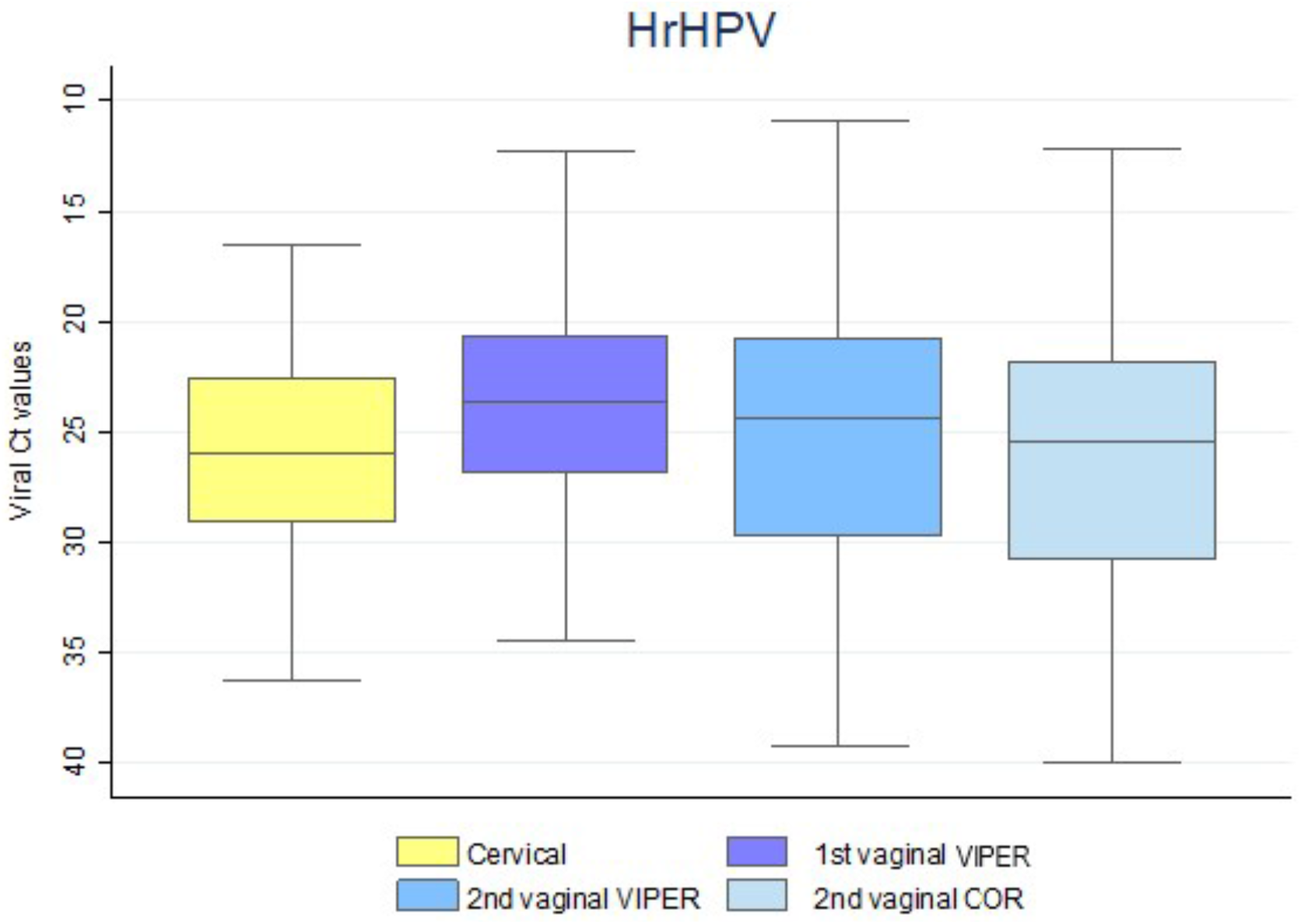
hrHPV cycle threshold (Ct) values for all sample types. In case of multiple infections, the lowest Ct value was considered. Boxplots indicate median Ct-values, interquartile ranges, and extreme values (whiskers).

## Discussion

In this study, we evaluated the clinical performance of FLOQSwab® vaginal self-collected samples analysed using the Onclarity assay on two dedicated automated platforms: the BD Viper™ and BD COR™. The findings show similar sensitivity for detection of ≥CIN2 and ≥CIN3 for the sequential self-collected vaginal specimens independent of analysis platform. With respect to the clinical performance of self-collected specimens and clinician collected routine samples, our data shows that the configuration of the BD Onclarity HPV Assay and FLOQSwab® collected self-samples is non-inferior to clinician-collected samples regarding clinical sensitivity. Moreover, overall and individual genotype concordance between vaginal self-samples and cervical samples varied between moderate and excellent (Table 3) similar to data already reported in the previous study (13). A very good agreement in hrHPV detection was observed comparing the results obtained from the 1st and the 2nd self-collected vaginal samples tested using BD Viper™ and BD COR™ even if the second vaginal self-sample was resuspended at IEO laboratory in Italy, frozen and shipped to the Molecular Pathology Laboratory, Denmark. Moreover, no effect of sample order was observed in hrHPV detection as also reported in previous studies (27, 28).

Specificity for ≤CIN1 on self-samples was lower than the on clinician collected cervical samples. This contrasts with the data observed in a previous study by Latsuzbaia et al. looking at clinical performance of the BD Onclarity HPV test on self-collected samples where specificity of self-sample was higher compared to cervical sample (13). Different specificity measurements obtained using the same HPV test can be explained by considering the pre-analytical workflow preceding the HPV test. In the Belgian VALHUDES study by Latsuzbaia et al., self-collected samples were resuspended into 20 mL PreservCyt fixating medium (Hologic)(13) from which a fixed 800µl is retrieved for analysis on the automated BD Viper™ and BD COR™ platforms operating the Onclarity assay. In this study, the FLOQSwabs® self-collected samples were directly resuspended into the BD Onclarity HPV self-collection Diluent tube which holds 3 ml of lytic diluent. The aspiration volume for molecular analysis is the same as defined by the analysis platforms. Hence, the volume difference between the two resuspension protocols generates a much more concentrated sample in our setting compared to Latsuzbaia et al. A more concentrated sample leads to more HPV detection which translates into lower specificity. Moreover, the difference between a fixating medium like PreservCyt and a lytic medium like the HPV self-collection Diluent Tube can also be speculated to impact the accessibility of analytical material in the resulting suspension. A large reporting from the Dutch screening program by Inturrisi et al. (29) showed a similar observation, though with poorer performance of self-collected samples in both specificity and sensitivity compared to clinician collected samples. The Dutch analysis used Ct scores to show the difference between self-collected samples (Rovers Evalyn brush plus 20 ml PreservCyt) and clinician collected samples (Cervex Brush plus 20 ml PreservCyt) and found clinician collected samples to be more concentrated by extension of the Ct analysis. Combined with the Belgian VALHUDES, it is hardly surprisingly that the use of 20 ml PreservCyt to suspend self-collected samples could be speculated to result in a low concentration sample diluted to a point bordering the sensitivity and specificity tolerances to which modern day HPV assays are calibrated, given that they are calibrated against clinician collected specimens. Combined with our, although smaller, study this jointly points out the importance of evaluating the end-to-end pre-analytical and analytical workflow in the validation of self-collected samples for use with HPV molecular assays. To this end, the importance of different self-sampling devices and the resulting amount of material collected remains largely undocumented, with only a few studies have reported on different devices and the resulting accuracy in hrHPV detection with a clinical end point (13, 28, 30, 31), or from the point of view of analytical stability (32).

A final perspective regard work-flow. Using both the BD COR™ and the BD Viper™ platforms, swab samples where the user has broken of the FLOQSwab^®^ into an empty collection tube and shipped it to the laboratory can be processed upon reception by simply inserting the tube into the instrument for analysis. Whereas the BD Viper™ platform requires the laboratory to add diluent, the BD COR™ platform can add the diluent on-board and resuspend the sample as part of the preanalytical workflow, thereby practically rendering the operationalization of HPV self-sample testing hands-free. As self-sampling for cervical cancer screening becomes a more widely used screening modality such laboratory automations will have value in reducing the number of staff interactions required for analysis.

One way to modulate the clinical specificity and sensitivity of different combinations of resuspension media and brush types is to conduct *in silico* HPV assay cut off optimization, which in our case resulted in specificity improvement (Table S2). However, for cut off optimizations to have a general applicability to any HPV assay, the decision base should also include similar data from the larger intended use population. Another open question is whether the determinant of clinical performance is mainly driven by the choice of collection device or the medium and/or the resuspension volume (12). Nevertheless, some obvious lessons can be learned from the field today, in that using large volume LBC or diluents for self-collected sample applications can influence clinical performance. By extension, 3 ml diluents as used here or the 10 ml SurePath could provide stronger clinical performance concordance between clinician collected and self-collected samples even if it comes at the expense of a slightly lower specificity. However, besides resuspension volume other parameters may determine accuracy (33) and therefore the use of established validated protocols is crucial.

## Conclusions

In conclusion, it is important to validate collection device in combination with hrHPV assay using specific pre-analytical and analytical protocols for testing self-collected samples to demonstrate that results are reproducible and that there is no loss in accuracy due to the different procedures of specimen collection and processing.

hrHPV testing using BD Onclarity HPV™ assay on vaginal self-collected FLOQSwab® 5E089N using two different analysis platforms, BD Viper™ and BD COR™, has similar clinical sensitivity to detect ≥CIN2 compared to testing on clinician taken cervical samples. However, lower clinical specificity was observed on the self-samples but after analytical cut-off optimization, relative specificities did not differ from unity.

## List of abbreviations

CI: confidence interval
CIN: cervical intraepithelial neoplasia
CIN1: cervical intraepithelial neoplasia grade 1
CIN2: cervical intraepithelial neoplasia grade 2
CIN3: cervical intraepithelial neoplasia grade 3
Ct: cycle threshold
HPV: Human papillomavirus
HR: high-risk
hrHPV: high-risk Human papillomavirus
LBC: liquid-based cytology
UniMib: University of Milano-Bicocca
VALGENT: Validation of HPV Genotyping Tests
VALHUDES: Validation of Human Papillomavirus Assays and Collection Devices for Self-samples and Urine Samples

## Authors’ Contributions

Principal investigator and conceptualization: CEC, JB, MA. Protocol development: CEC, JB, MA. Funding acquisition: CEC, JB, MA. Project administration: CEC, JB, MA. Enrolment of patients: ADI, RP. Data curation and formal analysis: AL, MM, MA, ADI, RP. Sample handling and Methodology: MM, HP, FB, AFP. Drafting original manuscript: MM. Critical review and editing of manuscript: CEC, JB, MA, AL, ADI, FB, AFP, RP, HP, MM.

## Acknowledgments

MA and AL were supported by the Horizon 2020 Framework Programme for Research and Innovation of the European Commission, through the RISCC Network (Grant No. 847845).

## Extended Valhudes Study Group

**Chiara Giubbi**, Department of Medicine and Surgery, University of Milano-Bicocca, Monza, Italy;

**Si Brask Sonne**, Molecular Pathology Laboratory, Department of Pathology, af. 134, Copenhagen University Hospital, AHH-Hvidovre Hospital, Hvidovre, Denmark;

**Emilie Korsgaard Andrease**, Molecular Pathology Laboratory, Department of Pathology, af. 134, Copenhagen University Hospital, AHH-Hvidovre Hospital, Hvidovre, Denmark;

**Silvia Martella**, Preventive Gynecology Unit, European Institute of Oncology IRCCS, Milan, Italy;

**Eleonora Petra Preti**, Preventive Gynecology Unit, European Institute of Oncology IRCCS, Milan, Italy,:

**Maria Elena Guerrieri**, Preventive Gynecology Unit, European Institute of Oncology IRCCS, Milan, Italy;

**Rita Passerini**, Division of Laboratory Medicine, European Institute of Oncology IRCCS, Milan, Italy;

**Narcisa Muresu**, Department of Medicine, Surgery and Pharmacy, University of Sassari, Sassari, Italy

**Illari Sechi**, Department of Medicine, Surgery and Pharmacy, University of Sassari, Sassari, Italy;

**Arianna Dettori**, Department of Medicine, Surgery and Pharmacy, University of Sassari, Sassari, Italy;

**Maria Eugenia Ghi**, U.O. Coordinamento Consultori Familiari, ASSL Sassari – ATS Sardegna, Sassari, Italy;

**Maria Paola Bagella**, U.O. Coordinamento Consultori Familiari, ASSL Sassari – ATS Sardegna, Sassari, Italy;

**Adriano Marrazzu**, U.O. Coordinamento Consultori Familiari, ASSL Sassari – ATS Sardegna, Sassari, Italy;

