## Supplementary Tables 1 - 5 for "Performance of BD Onclarity HPV assay on FLOQSwabs vaginal self-samples"

**Table S1.** Absolute sensitivity for CIN2+ and CIN3+ and specificity for <CIN2 of BD Onclarity™ HPV Assay

| Sample type | N | Sensitivity [95%CI] ≥CIN2 (%) | N | Sensitivity [95%CI] ≥CIN3 (%) | N | Specificity [95%CI] ≤CIN1 (%) |
|---|---|---|---|---|---|---|
| **LBC** | 74/83 | 89.2 [80.4-94.9] | 44/48 | 91.7 [80-97.7] | 89/206 | 43.2 [36.3-50.3] |
| **1st vaginal** | 75/83 | 90.3 [81.9-95.7] | 44/48 | 91.7 [80-97.7] | 74/204 | 36.3 [29.7-43.3] |
| **2nd vaginal (BD Viper™)** | 74/82 | 90.2 [81.7-95.7] | 43/47 | 91.5 [79.6-97.6] | 68/205 | 33.2 [26.8-40.1] |
| **2nd vaginal (BD COR™)** | 74/83 | 89.1 [80.4-94.9] | 43/48 | 89.6 [77.3-96.5] | 74/206 | 35.9 [29.4-42.9] |

LBC, liquid based cytology

**Table S2.** Relative sensitivity and specificity with optimised cut-offs on self-samples.

| | Relative sensitivity [95%CI] ≥CIN2 | Relative sensitivity [95%CI] ≥CIN3 | Relative specificity [95%CI] ≤CIN1 |
|---|---|---|---|
| *Manufacturer´s cut-offs: HPV16 Ct<=38.3, others Ct<=34.2* | | | |
| 1st vaginal | 1.01 [0.97-1.06] | 1.00 [0.94-1.06] | 0.83 [0.73-0.94] |
| *Optimised cut-offs: HPV16 Ct≤38.3, HPV18 Ct ≤34.2, others Ct <=31.5* | | | |
| 1st vaginal | 1.01 [0.97-1.06] | 1.00 [0.94-1.06] | 0.91 [0.81-1.02] |

**Table S3.** Relative accuracy of BD Onclarity HPV assay on vaginal self-samples.

| | Relative sensitivity [95%CI] CIN2+ | Relative sensitivity [95%CI] CIN3 | Relative specificity [95%CI] <CIN2 |
|---|---|---|---|
| **1st vaginal (BD Viper™)** vs **2nd vaginal (BD Viper™)**[1] | 1.00 (0.96-1.04) | 1.00 (0.94-1.07) | 0.93 (0.85-1.02) |
| **1st vaginal (BD Viper™)** vs **2nd vaginal (BD COR™)**[2] | 0.99 (0.96-1.01) | 0.98 (0.90-1.06) | 1.00 (0.92-1.09) |
| **2nd vaginal (BD Viper™)** vs **2nd vaginal (BD COR™)**[3] | 0.99 (0.96-1.01) | 0.98 (0.93-1.02). | 1.07 (1.00-1.12) |

CIN, cervical intraepithelial neoplasia.

[1]HPV testing using Viper platform on first self-sample was considered as a comparator test and on second self-sample as an index test.

[2]HPV testing using Viper platform on first self-sample was considered as a comparator test and on second self-sample tested on COR platform as an index test.

[3]HPV testing using Viper platform on second self-sample was considered as a comparator test and on second self-sample tested on COR platform as an index test.

**Tables S4:** HPV test concordance between cervical and other specimens, overall and by disease status

### a) First vaginal sample tested on VIPER vs cervical sample

| | HPV type | +/+ | +/- | -/+ | -/- | Concordance [%] | Kappa [95% CI] |
|---|---|---|---|---|---|---|---|
| **Total population (n=286)** | hrHPV | 181 | 7 | 23 | 75 | 89.5 | 0.758 (0.677 - 0.839) |
| | HPV16 | 72 | 0 | 10 | 204 | 96.5 | 0.911 (0.857 - 0.965) |
| | HPV18 | 5 | 2 | 6 | 273 | 97.2 | 0.542 (0.261 - 0.823) |
| | HPV31 | 33 | 6 | 9 | 238 | 94.8 | 0.784 (0.679 - 0.889) |
| | HPV45 | 9 | 0 | 4 | 273 | 98.6 | 0.811 (0.631 - 0.992) |
| | HPV51 | 12 | 1 | 9 | 264 | 96.5 | 0.688 (0.507 - 0.869) |
| | HPV52 | 22 | 0 | 9 | 254 | 96.8 | 0.813 (0.695 - 0.931) |
| | HPV33/58 | 23 | 0 | 3 | 260 | 99.0 | 0.933 (0.858 - 1.000) |
| | HPV35/39/68 | 28 | 2 | 15 | 241 | 94.1 | 0.734 (0.615 - 0.853) |
| | HPV56/59/66 | 48 | 2 | 16 | 220 | 93.7 | 0.804 (0.717 - 0.890) |
| | **HPV type** | **+/+** | **+/-** | **-/+** | **-/-** | **Concordance [%]** | **Kappa [95% CI]** |
| **CIN2+ (n=83)** | hrHPV | 73 | 1 | 2 | 7 | 96.4 | 0.803 (0.588 - 1.000) |
| | HPV16 | 43 | 0 | 4 | 36 | 95.2 | 0.903 (0.811 - 0.995) |
| | HPV18 | 4 | 1 | 0 | 78 | 98.8 | 0.883 (0.655 - 1.000) |
| | HPV31 | 18 | 0 | 0 | 65 | 100.0 | 1.000 (1.000 - 1.000) |
| | HPV45 | 3 | 0 | 3 | 77 | 96.4 | 0.650 (0.285 - 1.000) |
| | HPV51 | 1 | 0 | 3 | 79 | 96.4 | 0.388 (0.149 - 0.926) |
| | HPV52 | 6 | 0 | 3 | 74 | 96.4 | 0.781 (0.544 - 1.000) |
| | HPV33/58 | 12 | 0 | 1 | 70 | 98.8 | 0.953 (0.861 - 1.000) |
| | HPV35/39/68 | 7 | 1 | 7 | 68 | 90.4 | 0.586 (0.332 - 0.839) |
| | HPV56/59/66 | 10 | 0 | 3 | 70 | 96.4 | 0.849 (0.683 - 1.000) |
| | **HPV type** | **+/+** | **+/-** | **-/+** | **-/-** | **Concordance [%]** | **Kappa [95% CI]** |
| **<CIN2 (n=203)** | hrHPV | 108 | 6 | 21 | 68 | 86.7 | 0.725 (0.629 - 0.820) |
| | HPV16 | 29 | 0 | 6 | 168 | 97.0 | 0.889 (0.802 - 0.976) |
| | HPV18 | 1 | 1 | 6 | 195 | 96.6 | 0.210 (-0.150 - 0.570) |
| | HPV31 | 15 | 6 | 9 | 173 | 92.6 | 0.625 (0.452 - 0.799) |
| | HPV45 | 6 | 0 | 1 | 196 | 99.5 | 0.921 (0.766 - 1.000) |
| | HPV51 | 11 | 1 | 6 | 185 | 96.6 | 0.741 (0.558 - 0.924) |
| | HPV52 | 16 | 0 | 6 | 180 | 97.0 | 0.826 (0.691 - 0.961) |
| | HPV33/58 | 11 | 0 | 2 | 190 | 99.0 | 0.911 (0.790 - 1.000 |
| | HPV35/39/68 | 21 | 1 | 8 | 173 | 95.6 | 0.799 (0.672 - 0.925) |
| | HPV56/59/66 | 38 | 2 | 13 | 150 | 92.6 | 0.788 (0.687 - 0.890) |

N, number; CI, 95% confidence interval; CIN, cervical intraepithelial neoplasia

+/+ positive on 1st vaginal and cervical samples, +/- positive only on cervical samples, -/+ positive only on 1st vaginal sample, -/- negative on both sample types.

Note: 0.00 – 0.20 Poor; 0.21 – 0.40 Fair; 0.41 – 0.60 Moderate; 0.61 – 0.80 Good; 0.81 – 1.00 Excellent (3).

**b) Second vaginal sample tested with VIPER vs cervical samples**

| | HPV type | +/+ | +/- | -/+ | -/- | Concordance [%] | Kappa [95% CI] |
|---|---|---|---|---|---|---|---|
| **Total population (n=286)** | hrHPV | 184 | 4 | 26 | 72 | 89.5 | 0.754 (0.672 - 0.836) |
| | HPV16 | 72 | 0 | 14 | 200 | 95.1 | 0.878 (0.816 - 0.940) |
| | HPV18 | 5 | 2 | 4 | 275 | 97.9 | 0.614 (0.331 - 0.898) |
| | HPV31 | 36 | 3 | 12 | 235 | 94.8 | 0.797 (0.698 - 0.896) |
| | HPV45 | 9 | 0 | 3 | 274 | 99.0 | 0.852 (0.687 - 1.000) |
| | HPV51 | 13 | 0 | 5 | 268 | 98.3 | 0.830 (0.684 - 0.976) |
| | HPV52 | 22 | 0 | 6 | 258 | 97.9 | 0.869 (0.766 - 0.972) |
| | HPV33/58 | 22 | 1 | 5 | 258 | 97.9 | 0.869 (0.765 - 0.972) |
| | HPV35/39/68 | 30 | 0 | 13 | 243 | 95.5 | 0.797 (0.691 - 0.902) |
| | HPV56/59/66 | 49 | 1 | 15 | 221 | 94.4 | 0.825 (0.743 - 0.907) |
| | **HPV type** | **+/+** | **+/-** | **-/+** | **-/-** | **Concordance [%]** | **Kappa [95% CI]** |
| **CIN2+ (n=82)** | hrHPV | 72 | 1 | 2 | 7 | 96.3 | 0.803 (0.588 - 1.000) |
| | HPV16 | 42 | 0 | 3 | 37 | 96.3 | 0.927 (0.845 - 1.000) |
| | HPV18 | 4 | 1 | 0 | 77 | 98.8 | 0.883 (0.655 - 1.000) |
| | HPV31 | 18 | 0 | 0 | 64 | 100.0 | 1.000 (1.000 - 1.000) |
| | HPV45 | 3 | 0 | 2 | 77 | 97.6 | 0.738 (0.392 - 1.000) |
| | HPV51 | 1 | 0 | 2 | 79 | 97.6 | 0.491 (-0.109 - 1.000) |
| | HPV52 | 6 | 0 | 3 | 73 | 96.3 | 0.781 (0.543 - 1.000) |
| | HPV33/58 | 11 | 1 | 4 | 66 | 93.9 | 0.779 (0.594 - 0.964) |
| | HPV35/39/68 | 8 | 0 | 3 | 71 | 96.3 | 0.822 (0.627 - 1.000) |
| | HPV56/59/66 | 10 | 0 | 4 | 68 | 95.1 | 0.806 (0.624 - 0.988) |
| | **HPV type** | **+/+** | **+/-** | **-/+** | **-/-** | **Concordance [%]** | **Kappa [95% CI]** |
| **<CIN2 (n=204)** | hrHPV | 112 | 3 | 24 | 65 | 86.8 | 0.724 (0.629 - 0.818) |
| | HPV16 | 30 | 0 | 11 | 163 | 94.6 | 0.813 (0.708 - 0.919) |
| | HPV18 | 1 | 1 | 4 | 198 | 97.6 | 0.276 (-0.163 - 0.715) |
| | HPV31 | 18 | 3 | 12 | 171 | 92.7 | 0.665 (0.509 - 0.821) |
| | HPV45 | 6 | 0 | 1 | 197 | 99.5 | 0.921 (0.766 - 1.000) |
| | HPV51 | 12 | 0 | 3 | 189 | 98.5 | 0.881 (0.749 - 1.000) |
| | HPV52 | 16 | 0 | 3 | 185 | 98.5 | 0.906 (0.802 - 1.000) |
| | HPV33/58 | 11 | 0 | 1 | 192 | 99.1 | 0.954 (0.864 - 1.000) |
| | HPV35/39/68 | 22 | 0 | 10 | 172 | 95.1 | 0.788 (0.662 - 0.913) |
| | HPV56/59/66 | 39 | 1 | 11 | 153 | 94.1 | 0.830 (0.737 - 0.922) |

N, number; CI, 95% confidence interval; CIN, cervical intraepithelial neoplasia

+/+ positive on vaginal self- and cervical samples, +/- positive only on cervical samples, -/+ positive only on vaginal self-samples, -/- negative on both sample types.

Note: 0.00 – 0.20 Poor; 0.21 – 0.40 Fair; 0.41 – 0.60 Moderate; 0.61 – 0.80 Good; 0.81 – 1.00 Excellent (3).

#### c) Second vaginal sample tested with COR vs cervical samples

| | HPV type | +/+ | +/- | -/+ | -/- | Concordance [%] | Kappa [95% CI] |
|---|---|---|---|---|---|---|---|
| **Total population (n=288)** | hrHPV | 184 | 6 | 22 | 76 | 90.3 | 0.775 (0.696 - 0.853) |
| | HPV16 | 73 | 0 | 12 | 203 | 95.8 | 0.896 (0.838 - 0.953) |
| | HPV18 | 4 | 3 | 4 | 277 | 97.6 | 0.521 (0.210 - 0.832) |
| | HPV31 | 35 | 5 | 11 | 237 | 94.4 | 0.781 (0.679 - 0.884) |
| | HPV45 | 9 | 0 | 3 | 276 | 99.0 | 0.852 (0.687 - 1.000) |
| | HPV51 | 13 | 0 | 5 | 270 | 98.3 | 0.830 (0.684 - 0.976) |
| | HPV52 | 22 | 0 | 7 | 259 | 97.6 | 0.850 (0.741 - 0.958) |
| | HPV33/58 | 22 | 1 | 3 | 262 | 98.6 | 0.909 (0.821 - 0.997) |
| | HPV35/39/68 | 30 | 0 | 10 | 248 | 96.5 | 0.838 (0.740 - 0.935) |
| | HPV56/59/66 | 49 | 1 | 14 | 224 | 94.8 | 0.835 (0.755 - 0.916) |
| | **HPV type** | **+/+** | **+/-** | **-/+** | **-/-** | **Concordance [%]** | **Kappa [95% CI]** |
| **CIN2+ (n=83)** | hrHPV | 72 | 2 | 2 | 7 | 95.2 | 0.751 (0.517 - 0.984) |
| | HPV16 | 43 | 0 | 3 | 37 | 96.4 | 0.927 (0.847 - 1.000) |
| | HPV18 | 3 | 2 | 0 | 78 | 97.6 | 0.738 (0.392 - 1.000) |
| | HPV31 | 17 | 1 | 0 | 65 | 100.0 | 0.964 (0.893 - 1.000) |
| | HPV45 | 3 | 0 | 2 | 78 | 97.6 | 0.738 (0.392 - 1.000) |
| | HPV51 | 1 | 0 | 2 | 80 | 97.6 | 0.491 (-0.109 - 1.000) |
| | HPV52 | 6 | 0 | 3 | 74 | 96.4 | 0.781 (0.544 - 1.000) |
| | HPV33/58 | 11 | 1 | 2 | 69 | 96.4 | 0.859 (0.703 - 1.000 |
| | HPV35/39/68 | 8 | 0 | 3 | 72 | 96.4 | 0.822 (0.628 - 1.000) |
| | HPV56/59/66 | 10 | 0 | 3 | 70 | 96.4 | 0.849 (0.683 - 1.000) |
| | **HPV type** | **+/+** | **+/-** | **-/+** | **-/-** | **Concordance [%]** | **Kappa [95% CI]** |
| **<CIN2 (n=205)** | hrHPV | 112 | 4 | 20 | 69 | 88.3 | 0.757 (0.666 - 0.847) |
| | HPV16 | 30 | 0 | 9 | 166 | 95.6 | 0.844 (0.745 - 0.942) |
| | HPV18 | 1 | 1 | 4 | 199 | 97.6 | 0.276 (-0.163 - 0.715) |
| | HPV31 | 18 | 4 | 11 | 172 | 92.7 | 0.665 (0.508 - 0.822) |
| | HPV45 | 6 | 0 | 1 | 198 | 99.5 | 0.921 (0.766 - 1.000) |
| | HPV51 | 12 | 0 | 3 | 190 | 98.5 | 0.881 (0.749 - 1.000) |
| | HPV52 | 16 | 0 | 4 | 185 | 98.1 | 0.878 (0.761 - 0.996) |
| | HPV33/58 | 11 | 0 | 1 | 193 | 99.5 | 0.954 (0.864 - 1.000) |
| | HPV35/39/68 | 22 | 0 | 7 | 176 | 96.6 | 0.844 (0.731 - 0.956) |
| | HPV56/59/66 | 39 | 1 | 11 | 154 | 94.2 | 0.830 (0.737 - 0.922) |

N, number; CI, 95% confidence interval; CIN, cervical intraepithelial neoplasia
+/+ positive on vaginal self- and cervical samples, +/- positive only on cervical samples, -/+ positive only on vaginal self-samples, -/- negative on both sample types.
Note: 0.00 – 0.20 Poor; 0.21 – 0.40 Fair; 0.41 – 0.60 Moderate; 0.61 – 0.80 Good; 0.81 – 1.00 Excellent (3).

d) **First vaginal sample tested with VIPER vs second vaginal tested with VIPER**

| | HPV type | +/+ | +/- | -/+ | -/- | Concordance [%] | Kappa [95% CI] |
|---|---|---|---|---|---|---|---|
| **Total population (n=285)** | hrHPV | 200 | 4 | 9 | 72 | 95.4 | 0.886 (0.825 - 0.946) |
| | HPV16 | 80 | 1 | 5 | 199 | 97.9 | 0.949 (0.909 - 0.989) |
| | HPV18 | 8 | 3 | 1 | 273 | 98.6 | 0.793 (0.595 - 0.990) |
| | HPV31 | 42 | 0 | 5 | 238 | 98.3 | 0.933 (0.876 - 0.991) |
| | HPV45 | 12 | 1 | 0 | 272 | 99.7 | 0.958 (0.876 - 1.000) |
| | HPV51 | 16 | 5 | 2 | 262 | 97.5 | 0.807 (0.669 - 0.946) |
| | HPV52 | 27 | 5 | 1 | 252 | 97.9 | 0.888 (0.800 - 0.976) |
| | HPV33/58 | 24 | 2 | 3 | 256 | 98.3 | 0.896 (0.806 - 0.986) |
| | HPV35/39/68 | 39 | 5 | 5 | 236 | 96.5 | 0.866 (0.784 - 0.947) |
| | HPV56/59/66 | 60 | 4 | 4 | 217 | 97.2 | 0.919 (0.864 - 0.974) |
| | **HPV type** | **+/+** | **+/-** | **-/+** | **-/-** | **Concordance [%]** | **Kappa [95% CI]** |
| **CIN2+ (n=82)** | hrHPV | 73 | 1 | 1 | 7 | 97.6 | 0.861 (0.673 - 1.000) |
| | HPV16 | 45 | 1 | 0 | 36 | 98.8 | 0.975 (0.927 - 1.000) |
| | HPV18 | 3 | 1 | 1 | 77 | 97.6 | 0.737 (0.388 - 1.000) |
| | HPV31 | 18 | 0 | 0 | 64 | 100.0 | 1.000 (1.000 - 1.000) |
| | HPV45 | 5 | 1 | 0 | 76 | 98.8 | 0.903 (0.714 - 1.000) |
| | HPV51 | 2 | 2 | 1 | 77 | 96.3 | 0.553 (0.102 - 1.000) |
| | HPV52 | 9 | 0 | 0 | 73 | 100.0 | 1.000 (1.000 - 1.000) |
| | HPV33/58 | 12 | 1 | 3 | 66 | 95.1 | 0.828 (0.665 - 0.991) |
| | HPV35/39/68 | 10 | 4 | 1 | 67 | 93.9 | 0.765 (0.569 - 0.961) |
| | HPV56/59/66 | 13 | 0 | 1 | 68 | 98.8 | 0.956 (0.869 - 1.000) |
| | **HPV type** | **+/+** | **+/-** | **-/+** | **-/-** | **Concordance [%]** | **Kappa [95% CI]** |
| **<CIN2 (n=203)** | hrHPV | 127 | 3 | 8 | 65 | 94.6 | 0.881 (0.812 - 0.949) |
| | HPV16 | 35 | 0 | 5 | 163 | 97.5 | 0.918 (0.848 - 0.989) |
| | HPV18 | 5 | 2 | 0 | 196 | 99.0 | 0.828 (0.595 - 1.000) |
| | HPV31 | 24 | 0 | 5 | 174 | 97.5 | 0.892 (0.798 - 0.985) |
| | HPV45 | 7 | 0 | 0 | 196 | 100.0 | 1.000 (1.000 - 1.000) |
| | HPV51 | 14 | 3 | 1 | 185 | 98.0 | 0.864 (0.734 - 0.995) |
| | HPV52 | 18 | 5 | 1 | 179 | 97.0 | 0.841 (0.717 - 0.965) |
| | HPV33/58 | 12 | 1 | 0 | 190 | 99.5 | 0.957 (0.874 - 1.000) |
| | HPV35/39/68 | 29 | 1 | 4 | 169 | 97.5 | 0.906 (0.825 - 0.987) |
| | HPV56/59/66 | 47 | 4 | 3 | 149 | 96.6 | 0.908 (0.841 - 0.975) |

N, number; CI, 95% confidence interval; CIN, cervical intraepithelial neoplasia

+/+ positive on 1st vaginal and 2nd vaginal sample tested on VIPER, +/- positive only on 1st vaginal sample, -/+ positive only on 2nd vaginal sample, -/- negative on both sample types.

Note: 0.00 – 0.20 Poor; 0.21 – 0.40 Fair; 0.41 – 0.60 Moderate; 0.61 – 0.80 Good; 0.81 – 1.00 Excellent (3).

e) **First vaginal sample tested with VIPER vs second vaginal tested with COR**

| | HPV type | +/+ | +/- | -/+ | -/- | Concordance [%] | Kappa [95% CI] |
|---|---|---|---|---|---|---|---|
| **Total population (n=287)** | hrHPV | 199 | 6 | 5 | 77 | 96.2 | 0.906 (0.852 - 0.961) |
| | HPV16 | 81 | 1 | 3 | 202 | 98.6 | 0.966 (0.933 - 0.999) |
| | HPV18 | 8 | 3 | 0 | 276 | 99.0 | 0.837 (0.656 - 1.000) |
| | HPV31 | 40 | 2 | 5 | 240 | 97.6 | 0.905 (0.836 - 0.974) |
| | HPV45 | 12 | 1 | 0 | 274 | 99.7 | 0.958 (0.876 - 1.000) |
| | HPV51 | 16 | 5 | 2 | 264 | 97.6 | 0.808 (0.669 - 0.946) |
| | HPV52 | 28 | 4 | 1 | 254 | 98.3 | 0.908 (0.829 - 0.988) |
| | HPV33/58 | 24 | 2 | 1 | 260 | 99.0 | 0.935 (0.863 - 1.000) |
| | HPV35/39/68 | 38 | 6 | 2 | 241 | 97.2 | 0.888 (0.813 - 0.964) |
| | HPV56/59/66 | 60 | 4 | 3 | 220 | 97.6 | 0.929 (0.877 - 0.981) |
| | **HPV type** | **+/+** | **+/-** | **-/+** | **-/-** | **Concordance [%]** | **Kappa [95% CI]** |
| **CIN2+ (n=83)** | hrHPV | 74 | 1 | 0 | 8 | 98.8 | 0.934 (0.807 - 1.000) |
| | HPV16 | 46 | 1 | 0 | 36 | 98.8 | 0.976 (0.928 - 1.000) |
| | HPV18 | 3 | 1 | 0 | 79 | 98.8 | 0.851 (0.564 - 1.000) |
| | HPV31 | 17 | 1 | 0 | 65 | 98.8 | 0.964 (0.893 - 1.000) |
| | HPV45 | 5 | 1 | 0 | 77 | 98.8 | 0.903 (0.714 - 1.000) |
| | HPV51 | 2 | 2 | 1 | 78 | 96.4 | 0.553 (0.102 - 1.000) |
| | HPV52 | 9 | 0 | 0 | 74 | 100.0 | 1.000 (1.000 - 1.000) |
| | HPV33/58 | 12 | 1 | 1 | 69 | 97.6 | 0.909 (0.784 - 1.000) |
| | HPV35/39/68 | 10 | 4 | 1 | 68 | 94.0 | 0.765 (0.569 - 0.961) |
| | HPV56/59/66 | 13 | 0 | 0 | 70 | 100.0 | 1.000 (1.000 - 1.000) |
| | **HPV type** | **+/+** | **+/-** | **-/+** | **-/-** | **Concordance [%]** | **Kappa [95% CI]** |
| **<CIN2 (n=204)** | hrHPV | 125 | 5 | 5 | 69 | 95.1 | 0.894 (0.830 - 0.958) |
| | HPV16 | 35 | 0 | 3 | 166 | 98.5 | 0.950 (0.894 - 1.000) |
| | HPV18 | 5 | 2 | 0 | 197 | 99.0 | 0.828 (0.595 - 1.000) |
| | HPV31 | 23 | 1 | 5 | 175 | 97.1 | 0.868 (0.764 - 0.971 |
| | HPV45 | 7 | 0 | 0 | 197 | 100.0 | 1.000 (1.000 - 1.000) |
| | HPV51 | 14 | 3 | 1 | 186 | 98.0 | 0.864 (0.734 - 0.995) |
| | HPV52 | 19 | 4 | 1 | 180 | 97.6 | 0.870 (0.758 - 0.982) |
| | HPV33/58 | 12 | 1 | 0 | 191 | 99.5 | 0.957 (0.874 - 1.000) |
| | HPV35/39/68 | 28 | 2 | 1 | 173 | 98.5 | 0.941 (0.874 - 1.000) |
| | HPV56/59/66 | 47 | 4 | 3 | 150 | 96.6 | 0.908 (0.841 - 0.975) |

N, number; CI, 95% confidence interval; CIN, cervical intraepithelial neoplasia
+/+ positive on 1st vaginal and 2nd vaginal sample tested on COR, +/- positive only on 1st vaginal sample, -/+ positive only on 2nd vaginal sample, -/- negative on both sample types.
Note: 0.00 – 0.20 Poor; 0.21 – 0.40 Fair; 0.41 – 0.60 Moderate; 0.61 – 0.80 Good; 0.81 – 1.00 Excellent (3).

### f) Second vaginal sample tested with VIPER vs second vaginal tested with COR

| | HPV type | +/+ | +/- | -/+ | -/- | Concordance [%] | Kappa [95% CI] |
|---|---|---|---|---|---|---|---|
| **Total population (n=287)** | hrHPV | 204 | 7 | 1 | 75 | 97.2 | 0.930 (0.883 - 0.978) |
| | HPV16 | 83 | 1 | 3 | 200 | 98.6 | 0.967 (0.934 - 0.999) |
| | HPV18 | 8 | 1 | 0 | 278 | 99.7 | 0.939 (0.821 - 1.000) |
| | HPV31 | 46 | 2 | 0 | 239 | 99.3 | 0.975 (0.939 - 1.000) |
| | HPV45 | 12 | 0 | 0 | 275 | 100.0 | 1.000 (1.000 - 1.000) |
| | HPV51 | 18 | 0 | 0 | 269 | 100.0 | 1.000 (1.000 - 1.000) |
| | HPV52 | 28 | 0 | 1 | 258 | 99.7 | 0.981 (0.942 - 1.000) |
| | HPV33/58 | 25 | 2 | 0 | 260 | 99.3 | 0.958 (0.899 - 1.000) |
| | HPV35/39/68 | 40 | 4 | 0 | 243 | 98.6 | 0.944 (0.890 - 0.998) |
| | HPV56/59/66 | 62 | 2 | 1 | 222 | 99.0 | 0.970 (0.936 - 1.000) |
| | **HPV type** | **+/+** | **+/-** | **-/+** | **-/-** | **Concordance [%]** | **Kappa [95% CI]** |
| **CIN2+ (n=82)** | hrHPV | 73 | 1 | 0 | 8 | 98.8 | 0.934 (0.807 - 1.000) |
| | HPV16 | 45 | 0 | 0 | 37 | 100.0 | 1.000 (1.000 - 1.000) |
| | HPV18 | 3 | 1 | 0 | 78 | 98.8 | 0.851 (0.564 - 1.000) |
| | HPV31 | 17 | 1 | 0 | 64 | 98.8 | 0.964 (0.893 - 1.000) |
| | HPV45 | 5 | 0 | 0 | 77 | 100.0 | 1.000 (1.000 - 1.000) |
| | HPV51 | 3 | 0 | 0 | 79 | 100.0 | 1.000 (1.000 - 1.000) |
| | HPV52 | 9 | 0 | 0 | 73 | 100.0 | 1.000 (1.000 - 1.000) |
| | HPV33/58 | 13 | 2 | 0 | 67 | 97.6 | 0.914 (0.797 - 1.000) |
| | HPV35/39/68 | 11 | 0 | 0 | 71 | 100.0 | 1.000 (1.000 - 1.000) |
| | HPV56/59/66 | 13 | 1 | 0 | 68 | 98.8 | 0.956 (0.869 - 1.000) |
| | **HPV type** | **+/+** | **+/-** | **-/+** | **-/-** | **Concordance [%]** | **Kappa [95% CI]** |
| **<CIN2 (n=205)** | hrHPV | 131 | 6 | 1 | 67 | 96.6 | 0.924 (0.869 - 0.979) |
| | HPV16 | 38 | 1 | 3 | 163 | 98.1 | 0.938 (0.878 - 0.998) |
| | HPV18 | 5 | 0 | 0 | 200 | 100.0 | 1.000 (1.000 - 1.000) |
| | HPV31 | 29 | 1 | 0 | 175 | 99.5 | 0.980 (0.942 - 1.000) |
| | HPV45 | 7 | 0 | 0 | 198 | 100.0 | 1.000 (1.000 - 1.000) |
| | HPV51 | 15 | 0 | 0 | 190 | 100.0 | 1.000 (1.000 - 1.000) |
| | HPV52 | 19 | 0 | 1 | 185 | 99.5 | 0.972 (0.916 - 1.000) |
| | HPV33/58 | 12 | 0 | 0 | 193 | 100.0 | 1.000 (1.000 - 1.000) |
| | HPV35/39/68 | 29 | 4 | 0 | 172 | 98.1 | 0.924 (0.851 - 0.998) |
| | HPV56/59/66 | 49 | 1 | 1 | 154 | 99.0 | 0.974 (0.937 - 1.000) |

N, number; CI, 95% confidence interval; CIN, cervical intraepithelial neoplasia

+/+ positive on 2nd vaginal sample tested on VIPER and 2nd vaginal sample tested on COR, +/- positive only on 2nd vaginal sample tested on VIPER, -/+ positive only on 2nd vaginal sample tested on COR, -/- negative on both samples.

Note: 0.00 – 0.20 Poor; 0.21 – 0.40 Fair; 0.41 – 0.60 Moderate; 0.61 – 0.80 Good; 0.81 – 1.00 Excellent (3).

**Supplementary Table 5a.** Difference in median Ct values between matched cervical and first vaginal sample tested in Milan on VIPER platform.

| | n[a] | Median Ct cervical [IQR] | Median Ct 1st vaginal [IQR] | p-value |
|---|---|---|---|---|
| hrHPV[b] | 181 | 25.9 [22.6-28.8] | 23.0 [20.3-25.9] | 0.000 |
| HPV16 | 75 | 26.9 [23.8 -29.8] | 25.5 [21.6-28.4] | 0.000 |
| HPV18 | 7 | 32.0 [29.3-34.8] | 28.7 [25.5-33.6] | 0.176 |
| HPV31 | 37 | 26.9 [24.8-31.2] | 25.2 [22.3-28.5] | 0.007 |
| HPV45 | 13 | 32.8 [28.8-34.3] | 27.9 [25.5-31.6] | 0.006 |
| HPV51 | 18 | 30.1 [25.7-34.5] | 26.0 [20.9-32.6] | 0.002 |
| HPV52 | 26 | 26.7 [24.3- 30.3] | 22.9 [21.5-26.8] | 0.000 |
| HPV33/58 | 26 | 26.6 [23.4-30.4] | 24.2 [22.7-26.9] | 0.016 |
| HPV35/39/68 | 41 | 31.5 [26.6-34.8] | 26.4 [23.0 -29.8] | 0.000 |
| HPV56/59/66 | 52 | 25.4 [22.2-29.1] | 21.7 [19.3- 25.5] | 0.000 |

[a]number of matched samples.
[b]In case of multiple HPV infections only the HPV type with the lowest Ct value was considered.
IQR, interquartile range (25-75%); Ct, cycle number; n, number.
Mann-Whitney test was used to compare differences in median Ct values of matched cervical and 1st vaginal samples. Samples with Ct values above zero, including those beyond the cut-offs were considered for comparison.

**Supplementary Table 5b.** Difference in median Ct values between matched cervical and second vaginal sample tested in Hvidovre on VIPER platform.

| | n[a] | Median Ct cervical [IQR] | Median Ct 2nd vaginal VP [IQR] | p-value |
|---|---|---|---|---|
| hrHPV[b] | 188 | 25.9 [22.5-29.0] | 23.5 [20.1-26.5] | 0.000 |
| HPV16 | 75 | 26.9 [23.8 -29.8] | 26.1 [22.0-29.0] | 0.000 |
| HPV18 | 8 | 31.5 [29.8-33.1] | 31.2 [25.8-34.1] | 0.779 |
| HPV31 | 40 | 26.9 [24.3-31.3] | 26.0 [23.1-30.1] | 0.178 |
| HPV45 | 12 | 32.1 [28.2-34.0] | 26.9 [24.1-30.3] | 0.004 |
| HPV51 | 18 | 30.1 [25.7-34.5] | 26.5 [21.5-30.6] | 0.003 |
| HPV52 | 26 | 26.7 [24.3- 30.3] | 22.5 [20.1-26.4] | 0.000 |
| HPV33/58 | 26 | 26.6 [23.4-30.4] | 24.3 [22.1-28.7] | 0.101 |
| HPV35/39/68 | 42 | 31.4 [26.6-34.8] | 25.6 [21.9 -30.4] | 0.000 |
| HPV56/59/66 | 54 | 25.4 [22.0-29.4] | 21.6 [18.9- 25.5] | 0.000 |

[a]number of matched samples.
[b]In case of multiple HPV infections only the HPV type with the lowest Ct value was considered.
IQR, interquartile range (25-75%); Ct, cycle number; n, number.
Mann-Whitney test was used to compare differences in median Ct values of matched cervical and 2ndvaginal samples. Samples with Ct values above zero, including those beyond the cut-offs were considered for comparison.

**Supplementary Table 5c.** Difference in median Ct values between matched cervical and second vaginal sample tested in Hvidovre on COR platform.

| | n[a] | **Median Ct cervical [IQR]** | **Median Ct 2nd vaginal COR [IQR]** | **p-value** |
|---|---|---|---|---|
| hrHPV[b] | 190 | 25.9 [22.5-29.0] | 24.3 [21.2-27.6] | 0.000 |
| HPV16 | 76 | 26.7 [23.8 -29.7] | 27.1 [22.9-30.4] | 0.122 |
| HPV18 | 9 | 32.0 [30.2-33.4] | 33.1 [27.6-36.1] | 0.952 |
| HPV31 | 40 | 26.9 [24.3-31.3] | 27.1 [23.9-30.9] | 0.967 |
| HPV45 | 12 | 32.1 [28.2-34.0] | 28.4 [24.9-31.5] | 0.005 |
| HPV51 | 17 | 29.8 [25.7-32.3] | 26.8 [22.7-31.1] | 0.018 |
| HPV52 | 26 | 26.7 [24.3- 30.3] | 23.6 [21.4-27.5] | 0.000 |
| HPV33/58 | 25 | 26.3 [23.4-30.1] | 25.5 [23.2-29.3] | 0.241 |
| HPV35/39/68 | 42 | 31.4 [26.6-34.8] | 27.2 [23.5 -31.0] | 0.000 |
| HPV56/59/66 | 54 | 25.4 [22.0-29.4] | 21.6 [19.4- 26.3] | 0.000 |

[a]number of matched samples.
[b]In case of multiple HPV infections only the HPV type with the lowest Ct value was considered.
IQR, interquartile range (25-75%); Ct, cycle number; n, number.
Mann-Whitney test was used to compare differences in median Ct values of matched cervical and 2ndvaginal samples. Samples with Ct values above zero, including those beyond the cut-offs were considered for comparison.

**Supplementary Table 5d.** Difference in median Ct values between first vaginal sample tested in Milan on VIPER platform vs second vaginal sample tested in Hvidovre on VIPER platform.

| | n[a] | **Median Ct 1st vaginal VIP [IQR]** | **Median Ct 2nd vaginal VIP [IQR]** | **p-value** |
|---|---|---|---|---|
| hrHPV[b] | 203 | 23.6 [20.7-26.8] | 23.8 [20.3-27.2] | 0.216 |
| HPV16 | 82 | 25.9 [21.9-30.5] | 26.6 [22.4-31.2] | 0.125 |
| HPV18 | 13 | 32.4 [28.6-33.7] | 32.1 [27.3-36.9] | 0.700 |
| HPV31 | 50 | 26.2 [23.6-31.8] | 26.9 [23.7-31.3] | 0.484 |
| HPV45 | 15 | 27.9 [25.5-32.9] | 26.9 [24.9-31.8] | 0.088 |
| HPV51 | 30 | 31.3 [25.0-34.8] | 30.6 [24.0-36.3] | 0.205 |
| HPV52 | 34 | 25.3 [21.6-31.2] | 24.4 [20.8-32.4] | 0.596 |
| HPV33/58 | 28 | 24.3 [23.0-28.3] | 24.5 [22.3-30.0] | 0.158 |
| HPV35/39/68 | 50 | 27.8 [23.1 -31.2] | 26.8 [22.6 -33.1] | 0.685 |
| HPV56/59/66 | 69 | 23.7 [20.1- 29.2] | 23.8 [19.5- 29.6] | 0.586 |

[a]number of matched samples.
[b]In case of multiple HPV infections only the HPV type with the lowest Ct value was considered.
IQR, interquartile range (25-75%); Ct, cycle number; n, number.
Mann-Whitney test was used to compare differences in median Ct values of matched 1st vaginal and 2ndvaginal samples. Samples with Ct values above zero, including those beyond the cut-offs were considered for comparison.

**Supplementary Table 5e.** Difference in median Ct values between first vaginal sample tested in Milan on VIPER platform vs second vaginal sample tested in Hvidovre on COR platform.

| | n[a] | Median Ct 1st vaginal [IQR] | Median Ct 2nd vaginal COR [IQR] | p-value |
|---|---|---|---|---|
| hrHPV[b] | 203 | 23.8 [20.3-27.2] | 24.5 [21.4-28.4] | 0.000 |
| HPV16 | 83 | 26.6 [22.4-31.2] | 27.6 [23.0-32.3] | 0.000 |
| HPV18 | 12 | 32.1 [27.3-36.9] | 32.3 [27.5-36.5] | 0.084 |
| HPV31 | 48 | 26.9 [23.7-31.3] | 27.8 [24.2-32.4] | 0.002 |
| HPV45 | 15 | 26.9 [24.9-31.8] | 29.6 [26.0-33.0] | 0.348 |
| HPV51 | 27 | 30.6 [24.0-36.3] | 31.1 [25.1-36.7] | 0.003 |
| HPV52 | 33 | 24.4 [20.8-32.4] | 25.5 [22.1-31.1] | 0.001 |
| HPV33/58 | 27 | 24.5 [22.3-30.0] | 25.7 [23.2-30.4] | 0.004 |
| HPV35/39/68 | 49 | 26.8 [22.6 -33.1] | 27.6 [24.3 -33.7] | 0.000 |
| HPV56/59/66 | 66 | 23.8 [19.5- 29.6] | 24.2 [20.3- 30.2] | 0.001 |

[a]number of matched samples.

[b]In case of multiple HPV infections only the HPV type with the lowest Ct value was considered.

IQR, interquartile range (25-75%); Ct, cycle number; n, number.

Mann-Whitney test was used to compare differences in median Ct values of matched 1st and 2nd vaginal samples. Samples with Ct values above zero, including those beyond the cut-offs were considered for comparison.

**Supplementary Table 5f.** Difference in median Ct values between second vaginal sample tested in Hvidvore on VIPER platform vs second vaginal sample tested in Hvidovre on COR platform.

| | n[a] | Median Ct 2nd vaginal VP [IQR] | Median Ct 2nd vaginal COR [IQR] | p-value |
|---|---|---|---|---|
| hrHPV[b] | 224 | 24.1 [20.8-29.0] | 25.2 [21.6-30.2] | 0.000 |
| HPV16 | 85 | 26.6 [22.4-31.3] | 27.6 [23.0-32.3] | 0.000 |
| HPV18 | 15 | 32.6 [27.3-37.0] | 33.2 [27.6-38.2] | 0.002 |
| HPV31 | 54 | 27.2 [23.7-31.9] | 28.1 [24.2-33.1] | 0.000 |
| HPV45 | 15 | 26.8 [24.9-31.8] | 29.6 [26.0-33.0] | 0.004 |
| HPV51 | 30 | 30.6 [24.0-36.3] | 32.0 [25.1-37.1] | 0.000 |
| HPV52 | 36 | 24.7 [20.8-32.5] | 26.6 [22.4-32.3] | 0.000 |
| HPV33/58 | 29 | 24.8 [22.6-30.0] | 26.1 [23.5-30.9] | 0.000 |
| HPV35/39/68 | 54 | 27.2 [23.4 -33.6] | 28.5 [24.4 -34.9] | 0.000 |
| HPV56/59/66 | 75 | 24.7 [19.5- 30.7] | 25.5 [20.4- 31.0] | 0.000 |

[a]number of matched samples.

[b]In case of multiple HPV infections only the HPV type with the lowest Ct value was considered.

IQR, interquartile range (25-75%); Ct, cycle number; n, number.

Mann-Whitney test was used to compare differences in median Ct values of matched 2nd vaginal sample tested on COR and 2ndvaginal samples tested on VIPER. Samples with Ct values above zero, including those beyond the cut-offs were considered for comparison.
